# Trajectory of plasma lipidomes associated with the risk of late-onset Alzheimer’s disease pathogenesis: a longitudinal study in the ADNI cohort

**DOI:** 10.1101/2023.06.07.23291081

**Authors:** Tingting Wang, Matthias Arnold, Kevin Huynh, Patrick Weinisch, Corey Giles, Natalie A Mellett, Thy Duong, Bharadwaj Marella, Kwangsik Nho, Alysha De Livera, Xianlin Han, Colette Blach, Andrew J Saykin, Gabi Kastenmüller, Peter J Meikle, Rima Kaddurah-Daouk, the Alzheimer’s Disease Neuroimaging Initiative

## Abstract

Comprehensive lipidomic studies have demonstrated strong cross-sectional associations between the blood lipidome and late-onset Alzheimer’s disease (AD) and its risk factors. However, the longitudinal relationship between the lipidomic variations and progression of AD remains unknown. Here, we employed longitudinal lipidomic profiling on 4,730 plasma samples from 1,517 participants of the Alzheimer’s Disease Neuroimaging Initiative (ADNI) cohort to investigate the temporal evolution of lipidomes among diagnostic groups. At baseline, there were 1,393 participants including 437 cognitively normal (CN), 713 with mild cognitive impairment (MCI), and 243 AD cases. During follow up, 329 individuals (29 CN and 300 MCI) developed clinical AD (AD converters). We developed an AD-CN classification model to stratify the non-converting MCI group into AD-like and non AD-like MCI based on their lipidomics profiles at baseline. Longitudinal analysis identified associations between the change in ether lipid species (including alkylphosphatidylcholine, alkenylphosphatidylcholine, lysoalkylphosphatidylcholine, and lysoalkenylphosphatidylcholine) in converters relative to non-converting CN and MCI groups. Further, the AD-CN model efficiently classified MCI into low AD risk and high AD risk, with the high AD risk group having two times higher risk of conversion to AD than the low risk group. These findings suggest that the lipidomic profile can serve as a potential biomarker to identify individuals at higher risk for progressing to AD.

## Introduction

Late-onset Alzheimer’s disease (AD) is the leading cause of dementia, characterised by the progressive death of neurons and loss of brain structure, usually presenting with memory loss^1,2^. Many risk factors have been identified to collectively modulate risk for AD, with advanced age (≥ 65 years) being the strongest risk factor. Moreover, common genetic risk factors are associated with increased risk^3^, such as the *APOE ε*4 allele and sex, with females being more likely to develop AD (especially at age ≥80 years)^4^.

Spanning a period of 15-25 years, individuals with AD progress from cognitively normal(CN) through mild cognitive impairment (MCI) to overt dementia^2^. As a transitional state between CN and dementia, MCI has mixed aetiologies with different pathologies, neuropsychological profiles, or biomarker anomalies, often presenting with subtle to mild clinical symptoms^5,6^. The transition from MCI to dementia can take a varying length of time, with some individuals remaining stable or reverting to CN. The underlying molecular mechanisms contributing to this heterogeneity remain unknown. Accurate stratification of MCI using the molecular level information (such as lipidomic profiling) has the potential to improve the prognostic accuracy at the early stages of disease^7^, which is critical for streamlining clinical trials to shorten drug development cycle and avoid negative results due to this heterogeneity.

A growing number of studies have defined an intimate link between the plasma lipidome, measured at a single point in time, and AD^8-11^ or AD-related risk factors^12-17^. Besides, baseline lipid metabolic changes in AD patients were also demonstrated to be closely related with cerebrospinal fluid pathology markers, imaging features, and cognitive performance^18^. However, the plasma lipidome is highly dynamic, varying in response to environmental exposures (diet, physical activity)^19-21^ and over the longer term with age^22-25^. Similarly, the progression of cognitive impairment may predispose individuals to lifestyle changes (unbalanced diet or physical inactivity)^26,27^. These changes will influence peripheral lipid metabolism and may therefore appear to be associated with disease in a cross-sectional analysis (referred to as reverse causation). Longitudinal studies can minimise the impact of reverse causation by defining the relationship between changes in the plasma lipidome prior to AD, during the progression to AD, and in response to AD.

In this study, we performed longitudinal analysis of plasma lipidomic profiles in the Alzheimer’s Disease Neuroimaging Initiative (ADNI)-1, -GO and -2 cohorts to delineate the relationships between peripheral lipid metabolism and progression to AD. Using this complex data, we also assessed the utility of plasma lipids to identify MCI individuals at high risk of converting to AD.

## Results

For this study, we profiled a total of 4,730 longitudinal plasma samples from 1,517 participants of ADNI -1, -GO and -2 cohorts examined from baseline up to the 13^th^ time point (10^th^ years follow up period), with three major time points of baseline, 12 months, and 24 months that include the largest number of individuals (Figure 1). After quality control, the lipidomics profiles on all these measurements consist of 749 lipid species from 48 lipid classes (Supplementary Table 1).

**Figure 1.**
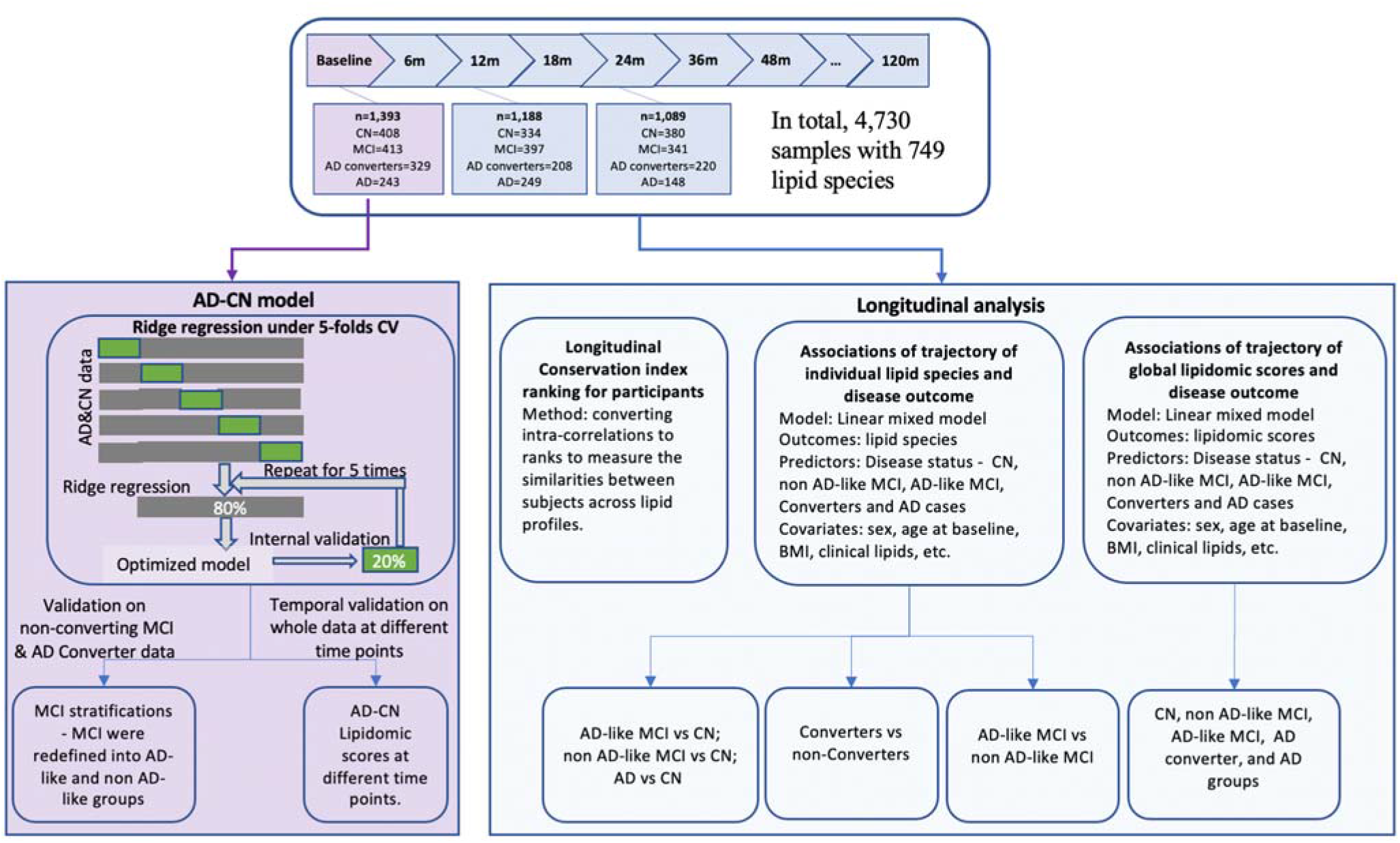
Study design. This study had three parts. Part 1 involved the development of an AD-CN risk model, using baseline data, to characterise the heterogeneity of the non-converting MCI group and to calculate lipidomic risk scores for individuals across different time points. A ridge regression model, built within a five-fold cross-validation framework was used to stratify the non-converting MCI group into AD-like and non AD-like sub-groups. In the development of the model, we treated AD-CN status as outcome with the predictors including all the lipid species, age, sex, BMI, and *APOE ε*4. Part 2 involved the calculation of a metabotype conservation index to quantify the stability of the lipidome over time. Part 3 was the longitudinal analysis on the repeated measurements across 13 time points to examine the associations of changes in lipid species and lipidomic risk scores with AD status. Associations of the trajectories of individual lipid species and disease outcomes were examined using linear mixed models to undercover the difference of trajectories of lipid species between different groups. The covariates included age, sex, BMI, HDL-C, triglycerides, cholesterol, fasting status, omega-3, and statin status. Thereafter, global lipidomic scores combing all lipid species derived from the AD-CN model were fitted into a linear mixed model to define the associations with disease outcomes. Similarly, the covariate set included age, sex, BMI, clinical lipids, fasting status, omega-3, and statin status.

### Longitudinal definition of diagnostic groups

We defined individuals as AD converters if they had a baseline diagnosis of cognitive normal (CN) or mild cognitive impairment (MCI) at the time that they entered the study but progressed to AD at a later time point. In total, there were 363 AD converters progressing from CN or MCI to AD at specific time points. In Supplementary Table 2, we detailed the distributions of AD converters across different time points. At baseline, after removing 25 missingness (detailed in the Method), we had 1,393 individuals with 437 CN, 713 MCI, and 243 AD cases. Out of these, there were 29 CN and 300 MCI who converted to AD at later time points (termed as AD converters). We also observed that 71 CN converted to MCI and remained MCI at subsequent time points, which we termed as MCI converters.

In Table 1, the difference in the demographics, numbers and distributions of the key risk factors (covariates) among diagnostic classifications were examined on three main time points (Baseline, 12 months, and 24 months), using Fisher’s exact test for categorical variables or ANOVA for continuous variables.

**Table 1.**
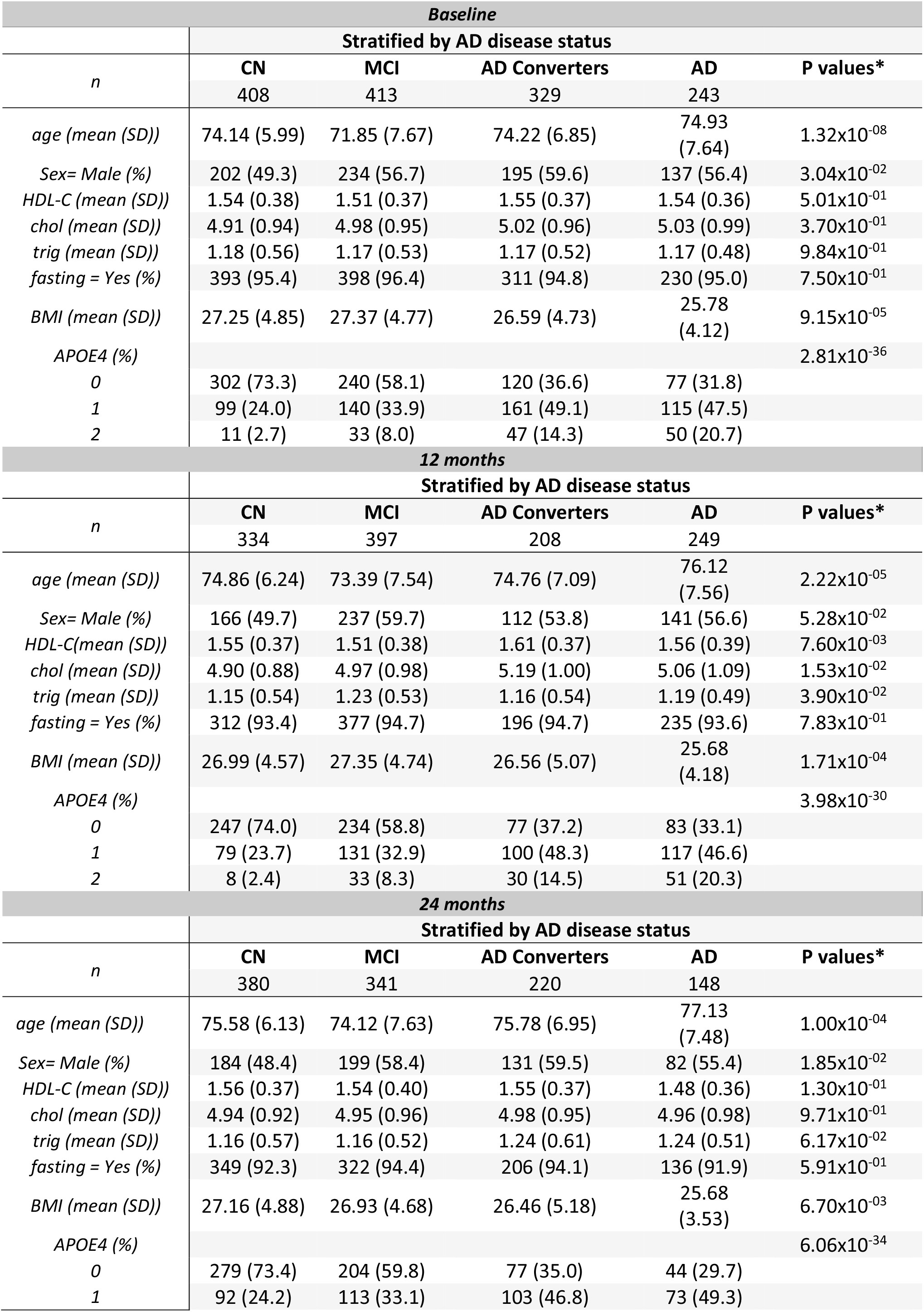
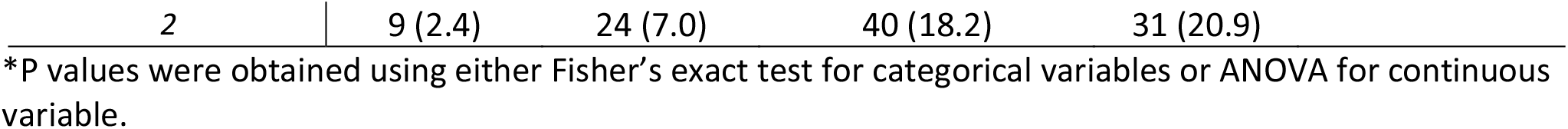
The basic characteristic of participants at Baseline, 12m and 24m.

| *Baseline* |  |  |  |  |  |
| --- | --- | --- | --- | --- | --- |
|  | **Stratified by AD disease status** |  |  |  |  |
| *n* | **CN** | **MCI** | **AD Converters** | **AD** | **P values\*** |
|  | 408 | 413 | 329 | 243 |  |
| *age (mean (SD))* | 74.14 (5.99) | 71.85 (7.67) | 74.22 (6.85) | 74.93 (7.64) | 1.32x10<sup>-08</sup> |
| *Sex= Male (%)* | 202 (49.3) | 234 (56.7) | 195 (59.6) | 137 (56.4) | 3.04x10<sup>-02</sup> |
| *HDL-C (mean (SD))* | 1.54 (0.38) | 1.51 (0.37) | 1.55 (0.37) | 1.54 (0.36) | 5.01x10<sup>-01</sup> |
| *chol (mean (SD))* | 4.91 (0.94) | 4.98 (0.95) | 5.02 (0.96) | 5.03 (0.99) | 3.70x10<sup>-01</sup> |
| *trig (mean (SD))* | 1.18 (0.56) | 1.17 (0.53) | 1.17 (0.52) | 1.17 (0.48) | 9.84x10<sup>-01</sup> |
| *fasting = Yes (%)* | 393 (95.4) | 398 (96.4) | 311 (94.8) | 230 (95.0) | 7.50x10<sup>-01</sup> |
| *BMI (mean (SD))* | 27.25 (4.85) | 27.37 (4.77) | 26.59 (4.73) | 25.78 (4.12) | 9.15x10<sup>-05</sup> |
| *APOE4 (%)* |  |  |  |  | 2.81x10<sup>-36</sup> |
| 0 | 302 (73.3) | 240 (58.1) | 120 (36.6) | 77 (31.8) |  |
| 1 | 99 (24.0) | 140 (33.9) | 161 (49.1) | 115 (47.5) |  |
| 2 | 11 (2.7) | 33 (8.0) | 47 (14.3) | 50 (20.7) |  |
| *12 months* |  |  |  |  |  |
|  | **Stratified by AD disease status** |  |  |  |  |
| *n* | **CN** | **MCI** | **AD Converters** | **AD** | **P values\*** |
|  | 334 | 397 | 208 | 249 |  |
| *age (mean (SD))* | 74.86 (6.24) | 73.39 (7.54) | 74.76 (7.09) | 76.12 (7.56) | 2.22x10<sup>-05</sup> |
| *Sex= Male (%)* | 166 (49.7) | 237 (59.7) | 112 (53.8) | 141 (56.6) | 5.28x10<sup>-02</sup> |
| *HDL-C (mean (SD))* | 1.55 (0.37) | 1.51 (0.38) | 1.61 (0.37) | 1.56 (0.39) | 7.60x10<sup>-03</sup> |
| *chol (mean (SD))* | 4.90 (0.88) | 4.97 (0.98) | 5.19 (1.00) | 5.06 (1.09) | 1.53x10<sup>-02</sup> |
| *trig (mean (SD))* | 1.15 (0.54) | 1.23 (0.53) | 1.16 (0.54) | 1.19 (0.49) | 3.90x10<sup>-02</sup> |
| *fasting = Yes (%)* | 312 (93.4) | 377 (94.7) | 196 (94.7) | 235 (93.6) | 7.83x10<sup>-01</sup> |
| *BMI (mean (SD))* | 26.99 (4.57) | 27.35 (4.74) | 26.56 (5.07) | 25.68 (4.18) | 1.71x10<sup>-04</sup> |
| *APOE4 (%)* |  |  |  |  | 3.98x10<sup>-30</sup> |
| 0 | 247 (74.0) | 234 (58.8) | 77 (37.2) | 83 (33.1) |  |
| 1 | 79 (23.7) | 131 (32.9) | 100 (48.3) | 117 (46.6) |  |
| 2 | 8 (2.4) | 33 (8.3) | 30 (14.5) | 51 (20.3) |  |
| *24 months* |  |  |  |  |  |
|  | **Stratified by AD disease status** |  |  |  |  |
| *n* | **CN** | **MCI** | **AD Converters** | **AD** | **P values\*** |
|  | 380 | 341 | 220 | 148 |  |
| *age (mean (SD))* | 75.58 (6.13) | 74.12 (7.63) | 75.78 (6.95) | 77.13 (7.48) | 1.00x10<sup>-04</sup> |
| *Sex= Male (%)* | 184 (48.4) | 199 (58.4) | 131 (59.5) | 82 (55.4) | 1.85x10<sup>-02</sup> |
| *HDL-C (mean (SD))* | 1.56 (0.37) | 1.54 (0.40) | 1.55 (0.37) | 1.48 (0.36) | 1.30x10<sup>-01</sup> |
| *chol (mean (SD))* | 4.94 (0.92) | 4.95 (0.96) | 4.98 (0.95) | 4.96 (0.98) | 9.71x10<sup>-01</sup> |
| *trig (mean (SD))* | 1.16 (0.57) | 1.16 (0.52) | 1.24 (0.61) | 1.24 (0.51) | 6.17x10<sup>-02</sup> |
| *fasting = Yes (%)* | 349 (92.3) | 322 (94.4) | 206 (94.1) | 136 (91.9) | 5.91x10<sup>-01</sup> |
| *BMI (mean (SD))* | 27.16 (4.88) | 26.93 (4.68) | 26.46 (5.18) | 25.68 (3.53) | 6.70x10<sup>-03</sup> |
| *APOE4 (%)* |  |  |  |  | 6.06x10<sup>-34</sup> |
| 0 | 279 (73.4) | 204 (59.8) | 77 (35.0) | 44 (29.7) |  |
| 1 | 92 (24.2) | 113 (33.1) | 103 (46.8) | 73 (49.3) |  |

\*P values were obtained using either Fisher’s exact test for categorical variables or ANOVA for continuous variable.

### Metabotype conservation over time among AD state groups

To globally assess changes in lipidomic profiles over time, we calculated metabotype conservation indices (*I*_*c*_) for a subset of 755 participants that had lipidomics data available at baseline as well as at follow up visits after 12 and 24 months. The *I*_*c*_ reflects the lipidomics-based self-similarity of an individual over time compared to all other individuals. To avoid potential bias introduced by the strong correlation structure observed in lipidomics data, we clustered highly correlated lipids and calculated eigenlipids for each of the 205 obtained clusters. Eighty lipids were not assigned to a cluster and retained as separate variables. Based on these 285 variables, we calculated the *I*_*c*_. As expected, the *I*_*c*_ after 24 months was overall significantly lower than after 12 months (paired t-test p = 1.06 × 10^−3^, Wilcoxon test p = 4.56 × 10^−6^); Figure 2 and Supplementary Table 3). *I*_*c*_ between baseline and 24-month follow-up showed overall high stability of the lipidome, with more than half of individuals reaching the maximum *I*_*c*_ of 1 (n = 401). There was no significant difference in the proportion of individuals with an *I*_*c*_ < 1 between diagnostic groups (Supplementary Table 4). However, comparing distributions of the *I*_*c*_ values smaller than one between diagnostic groups showed the highest conservation in the CN group, with significantly lower levels (Wilcoxon test p = 0.0039) observed in cases with AD (Figure 2b).

**Figure 2.**
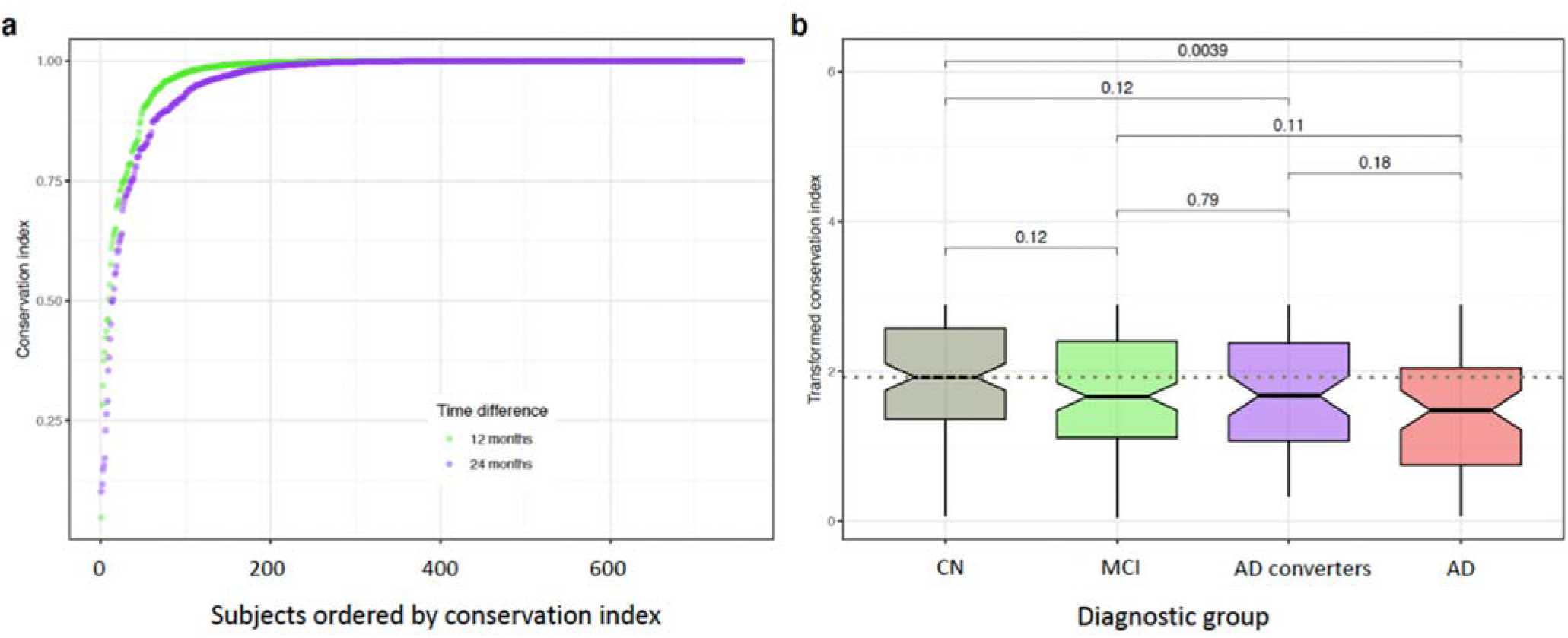
Metabotype conservation index. **a**. Comparison of metabotype conservation index (*I*_*c*_) across two time intervals (baseline to 12 and 24 months, respectively). **b**. Boxplot comparing the distribution of *I*_*c*_ values < 1 between diagnostic groups after 24 months. P-values are calculated using the Wilcoxon rank sum test and show a significantly lower *I*_*c*_ in the AD group compared to the CN group. For visualization purposes, we transformed the index using -log_10_(1 -*I*_*c*_).

### Consistent cross-sectional associations of individual lipid species with AD at different time points

At baseline, we observed 192 nominally significant associations between lipid species and AD, relative to CN, after FDR corrections (Figure 3, Supplementary Table 5). The majority of these associations were consistent with previous findings^28^. Using the 24-month data, there were 181 significant associations, whereas only 52 lipid species were significantly associated with the 12-month data (Figure 3). Among 192 associations identified at baseline, we observed that all of these associations were in the same direction as the ones identified at 24 months and only 2 out of 192 were different from the directions at 12 months. Pearson correlations between the beta coefficients (the associations between lipid species and AD state) among three time points were generally quite large (0.78 – 0.84).

**Figure 3.**
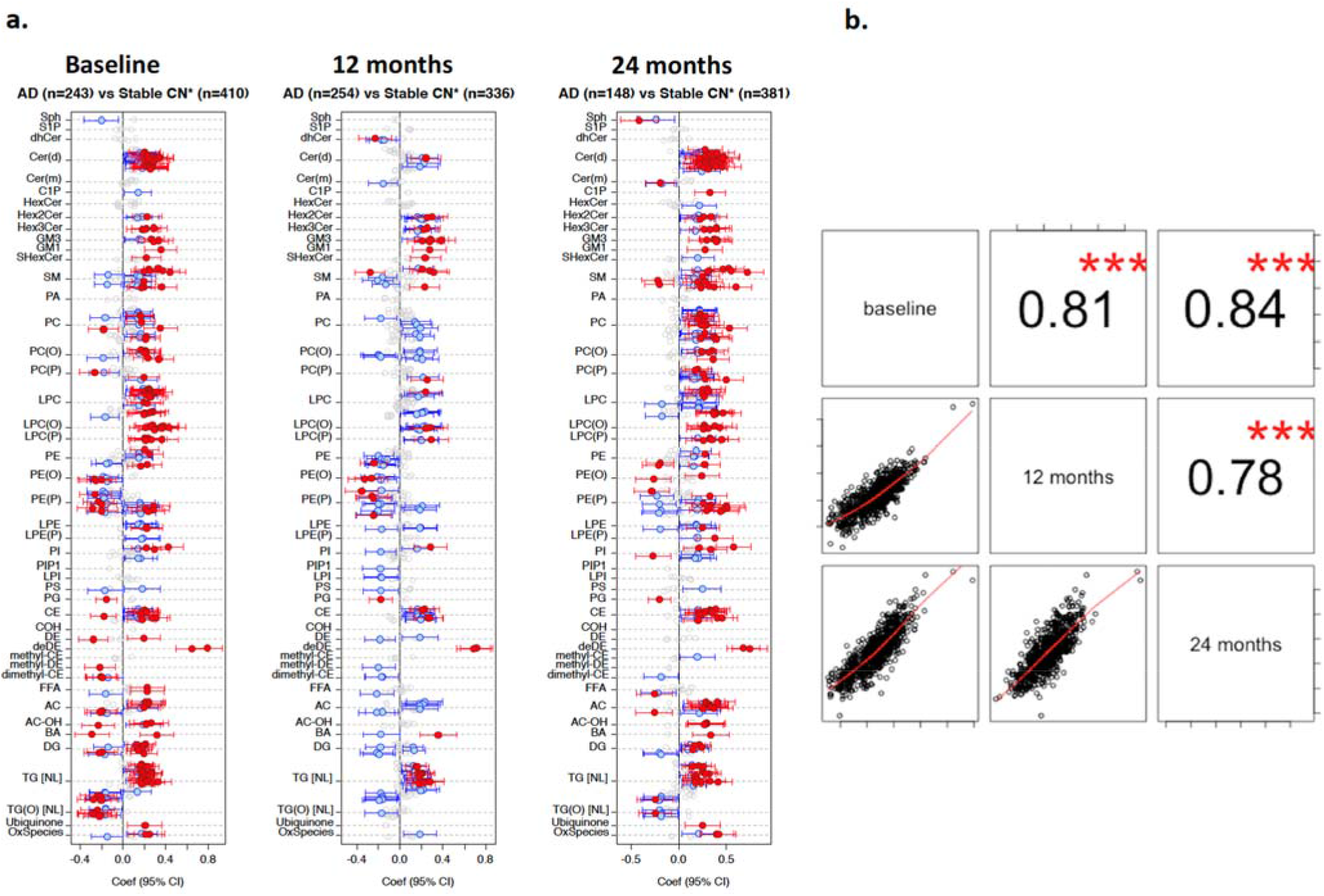
The associations of lipid species with AD state (AD vs CN). **a**. The associations of lipid species with AD versus CNl at different time points; **b**. Pearson correlations of beta coefficients among baseline, 12 months, and 24 months. Linear regression models of lipid species against AD adjusted for age, sex, BMI, clinical lipids, fasting status, cohort, omega-3, and statin status were performed on each time point (baseline, 12 month and 24 months). The coefficients were exacted from plot **a** and compared in the scatter plot **b**. Grey dots = not significant; blue dots = uncorrected p < 0.05; red dots = BH corrected p < 0.05, whiskers showed 95% confidence intervals.

Across the three time points, 35 lipid species were consistently associated with AD status. These lipid species originated from the ceramide (Cer(d)), dihexosylceramide (Hex2Cer), trihexosylceramide (Hex3Cer), GM3 ganglioside (GM3), sphingomyelin (SM), lysophosphatidylcholine (LPC), lysoalkylphosphatidylcholine (LPC(O)), lysoalkenylphosphatidylcholine (LPC(P)), alkylphosphatidylethanolamine (PE(O)), phosphatidylinositol (PI). There were two strongest associations observed between two novel lipid species from the dehydrodesmosterol ester (deDE) class, and AD across all three time points. In detail, they were deDE(18:2) (p=7.02×10^−25^ at baseline; 1.06×10^−17^ at 12 months; 8.80×10^−12^ at 24 months) and deDE(20:4) (p=7.48×10^−14^ at baseline; 2.32×10^−14^ at 12 months; 2.79×10^−12^ at 24 months). As shown in Supplementary Table 6, t-test results showed anticholinesterase medication usage was significantly different between AD and CN groups (0.0 % vs 87.2 %; CN vs AD; p-value<1.0×10^−04^). In the later analysis, we identified that both deDE lipid species showed the strongest associations with anticholinesterase medication at baseline (p=7.13×10^−18^ for deDE(18:2), p= 9.68×10^−12^ for deDE(20:4); Supplementary Table 7), indicating that they might be driven by the anticholinesterase medication usage (AD-related medication).

### The AD-CN model classified AD and stratified the non-converting MCI group into AD-like and non AD-like

An AD-CN model was developed using a ridge regression model within a 5-fold cross-validation framework on the AD and CN sub-cohort at baseline using all the lipid species (except the two deDE lipid species which were affected by the anticholinesterase medication usage) to predict AD status. The AD-CN model was applied to baseline profiles of individuals in the non-converting MCI and AD converter groups (the combinations of individuals converted from both MCI and CN groups; treated as true positives). The model could classify non-converting MCI and AD converters with an AUC of 0.69 (Figure 4). Relative to the basic model using four predictors – age, sex, BMI, and *APOE ε*4 (AUC=0.65; Supplementary Figure 1), our AD-CN model using the linear combination of the whole lipidome, age, sex, BMI, and APOE *ε*4 as the predictors showed better predictive power.

**Figure 4.**
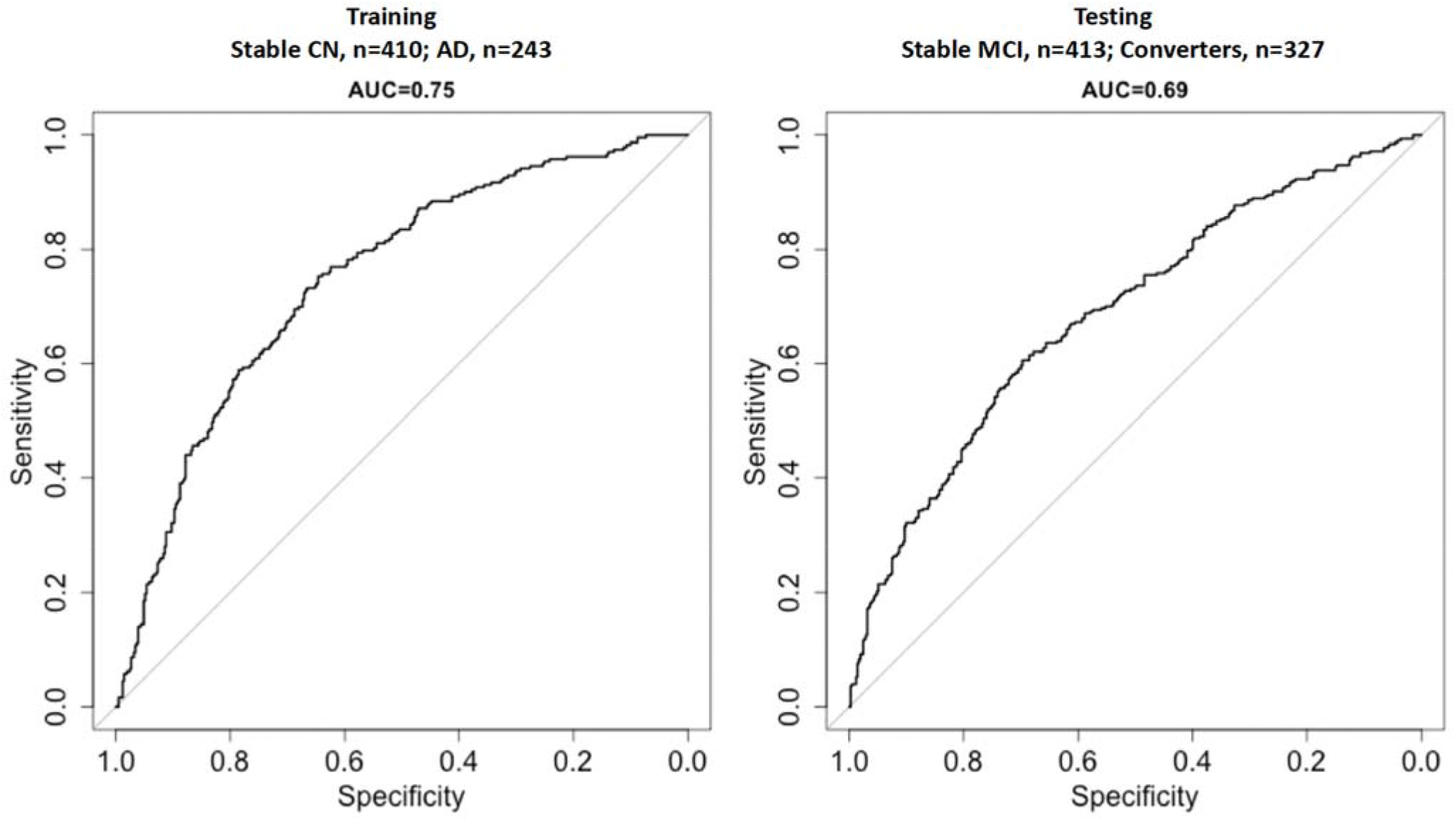
Prediction performance on training set (left) and testing set (right). Ridge regression model under 5-folds CV framework was performed using the disease status (training set -AD and CN group) as the binary outcome with a list of covariates including age, sex, BMI, fasting status, clinical lipids, cohort, omega-3, and statin status. The models were validated on non-converting MCI and converter groups with converters treated as true positives.

The predicted values from the model indicate the individual’s risk of developing AD. Using a cut-off point of 0.37 (illustrated in Supplementary Figure 2), the whole group (the combination of non-converting MCI and AD converters) was divided into low and high AD risk groups. The Kaplan-Meier plot (Figure 5a and 5b) showed the proportion of participants converted to AD in the high AD risk group across different time points was higher than in the low AD risk group. The Fisher’s exact test (p=4.85 ×10^−15^, Odd Ratio=3.14, 95% CI=2.30-4.31) also demonstrated that the model could efficiently stratify the group with most AD converters enriched in the high AD risk subgroup (59%).

**Figure 5.**
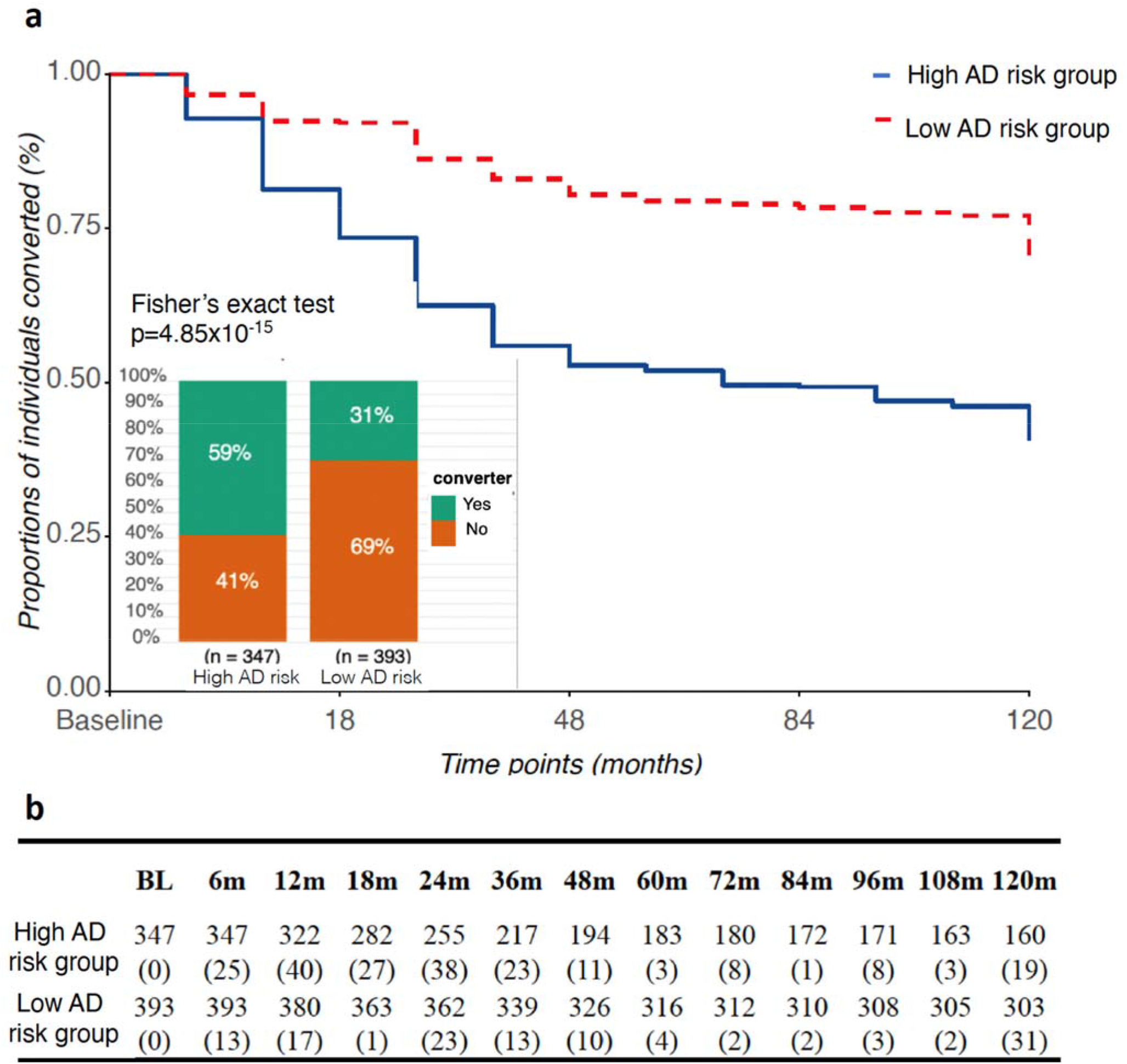
The proportion of converters in stratified non-converting MCI and converter groups. **a**. The proportions of individuals converted to AD at different time points between high AD and low AD risk groups (combination of non-converting and AD converters) were plotted in the Kaplan-Meier plot. The fisher’s exact test showed the distribution of converters in the high and low AD risk groups. **b**. The exact numbers of converters out of the total number of individuals at each time point were detailed in brackets.

Among 432 non-converting MCI, we further defined 271 individuals in the low AD risk group as non AD-like MCI and 142 individuals in the high AD risk group as AD-like MCI. We investigated the distribution of selected medications between the two non-converting MCI groups (Supplementary Table 8). Usage of Omega-3 was greater in the non-AD like MCI group (20.4 % vs 36.9 %; p =9.0×10^−04^), while anticholinesterase usage was greater in the AD-like MCI group (35.9 % vs 17.0 %; p<1.0×10^−04^).

### The performance of AD-CN model validated across the time points was stable

We further applied the AD-CN model built on the baseline AD and CN data to the whole data set (excluding AD converters) across different time points. To evaluate the prediction performance of the model on each single time point, we re-defined the AD diagnosis status of each individual as a binary variable – AD and non-AD status (combination of CN and MCI). The predictive performance at the three main time points was AUC: 0.71 (0.68 – 0.74) at baseline, AUC: 0.68 (0.64 – 0.71) at 12 months, and AUC: 0.72 (0.68 – 0.77) at 24 months (Figure 6). The model demonstrated consistent and robust prediction performance at each time point.

**Figure 6.**
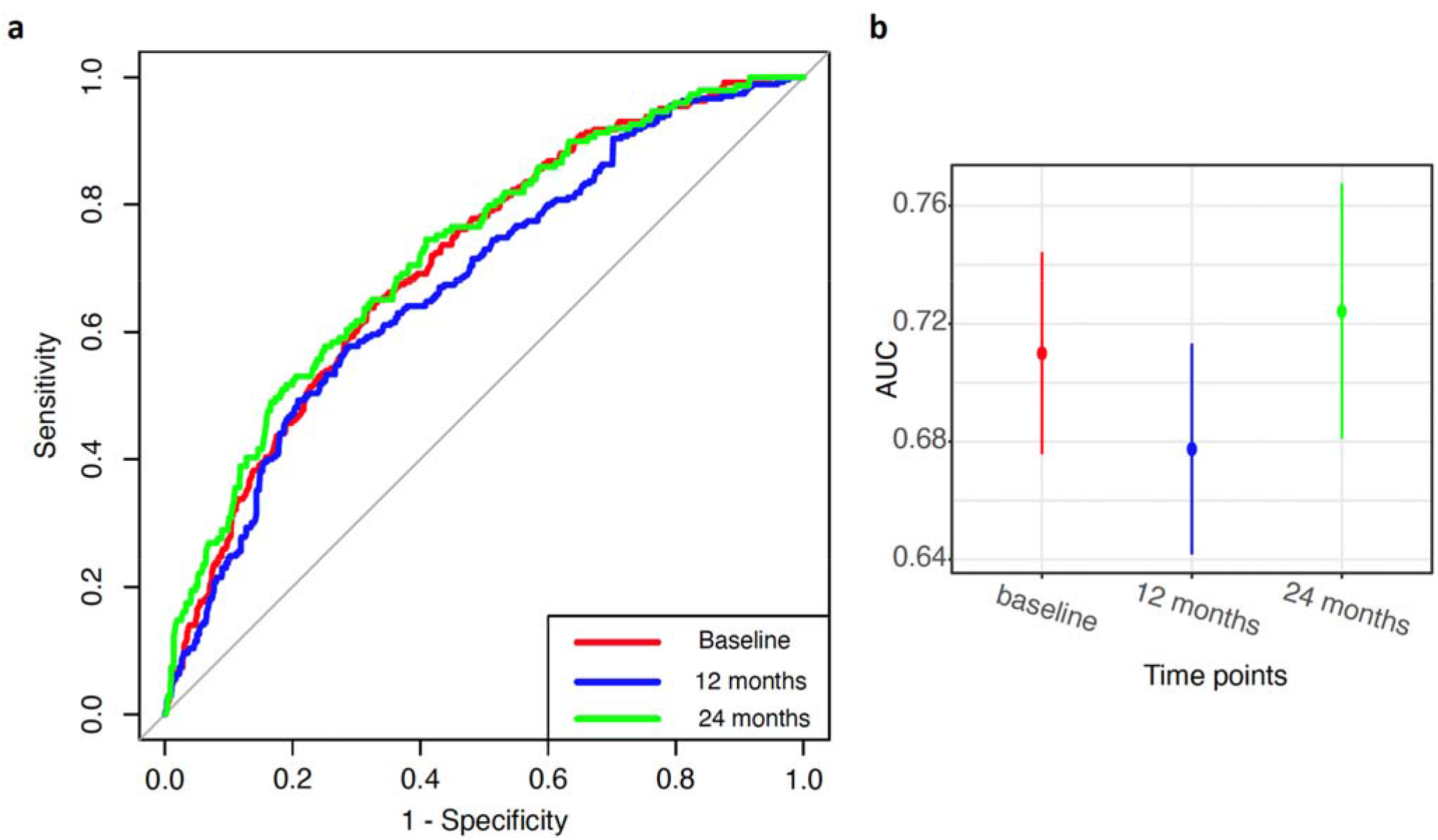
The performance of Lipidomic scores from AD-CN model across baseline, 12 months), and 24 months. a ROC AUC curves across baseline (red), 12 months (blue), and 24 months (green); b The AUC and 95% confident interval across baseline (red), 12 months (blue), and 24 months (green). The weights were first generated from ridge regression models on AD-CN data set at baseline using five-folds cross-validation. Then, the weights were separately applied to baseline, 12 months, and 24 months to generate the lipidomic scores at different time points. Further, the AUC was calculated on the lipidomic scores and the AD diagnosis status of each individuals (binary variable -AD and non-AD status that was the combination of CN and MCI).

### Trajectories of individual lipid species

The trajectories of individual lipid species were assessed using separate linear mixed-effects models. Significant interactions between AD status and time were observed for 1) the AD converter group versus the non-converter group (combination of non-converting CN and MCI); 2) AD-like versus non-AD-like MCI groups after excluding AD converters; and 3) non AD-like MCI vs CN after excluding AD converters.

When comparing AD and CN, only two lipid species (from the sphingosine class) were significant following FDR correction (Figure 7a; Supplementary Table 9).

**Figure 7.**
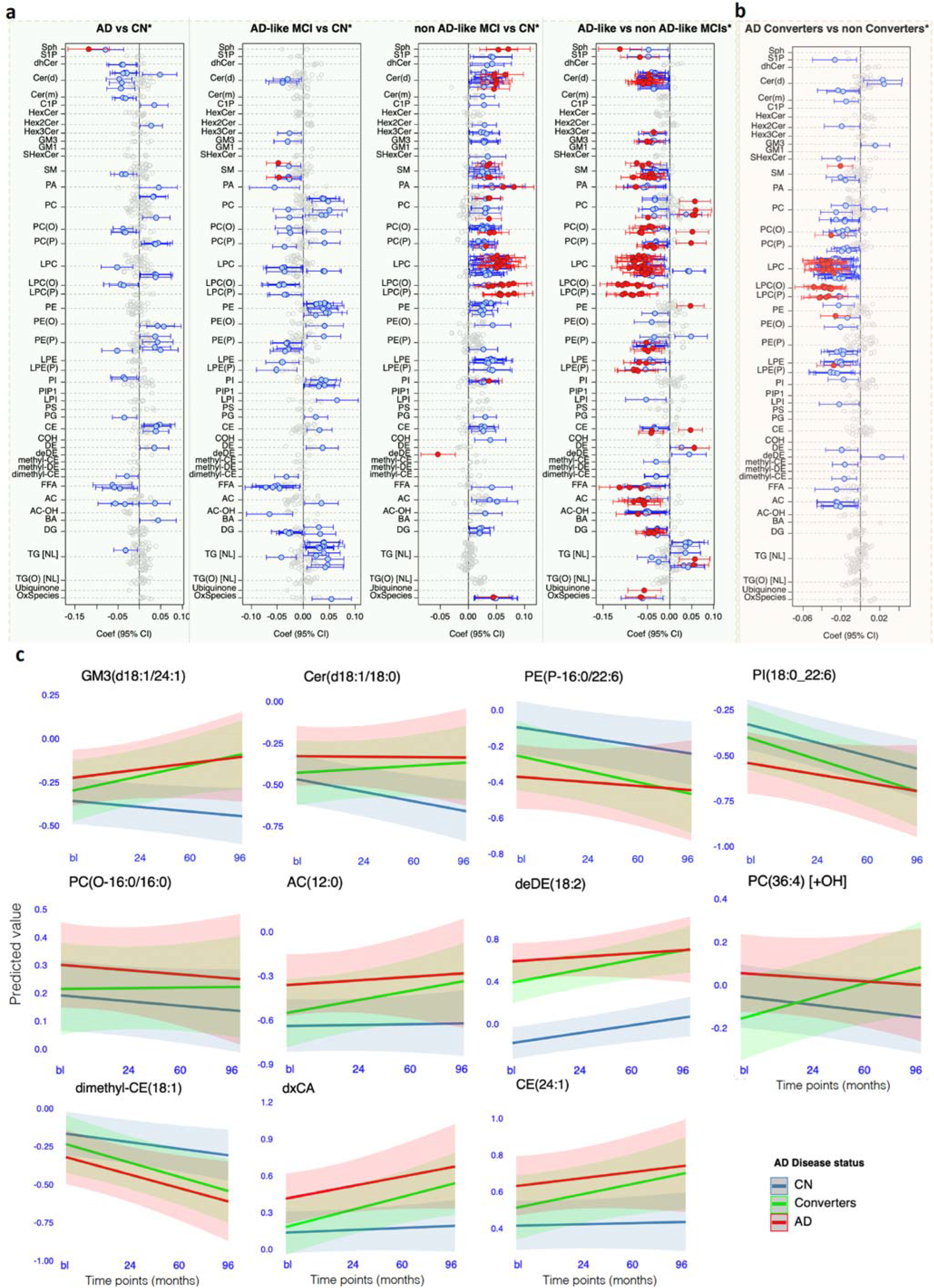
Trajectory of lipid species between different AD diagnosis groups. The linear mixed model was performed to examine the association of the changes of individual lipid species with AD diagnosis state: **a**. After excluding the AD converters, we compared trajectory of lipid species between the AD (652) vs non-converting CN (1,049), AD-like MCI (433) vs non-converting CN (1,049), and non AD-like MCI (781) vs non-converting CN (1,037); **b**. Using the similar model in the data sets excluding AD cases, we compared the converters (1,353) vs non-converter groups (the combination of non-converting CN and two non-converting MCI groups; 2,283). The covariates included age, sex, BMI, fasting status, HDL-C, total cholesterol, triglycerides, cohort, omega-3, and statin status; **c**. The trajectory of selected individual lipid species was further examined among the diagnosis groups of non-converting CN, AD converters, and AD using the above linear mixed model on the data set excluding MCI.

Comparing AD-like and non AD-like MCI groups, more than 121 lipid species from 26 lipid classes showed signficantly different trajectories after FDR correction (Figure 7a; Supplementary Table 9). A majority of these were from the SM, GM3, acylcarnitine (AC), alkylphosphatidylcholine (PC(O)), alkenylphosphatidylcholine (PC(P)), LPC, LPC(O), LPC(P), and dehydrocholesterol ester (DE) classes. Of these lipid species, we observed 13 lipid species from the lipid classes of GM3, LPC(O), AC, SM, CE and DE also appeared in the top 50 predictors in the AD-CN model.

The trajectories for 33 lipid species (after FDR correction) were found to significantly differ between AD converters vs non-converters (non-converting CN and MCI) (Figure 7b, Supplementary table 10). These lipids were primarily composed of the LPC(O), LPC(P), and PC(O) classes. The trajectory for all these lipid species were diametrically opposed with those between AD converters and non-converters. As a sensitivity analysis, we grouped MCI converters with AD converters and evaluated the associations of the trajectories of lipid species with the combination of AD and MCI converters (Supplementary Figure 3, Supplementary table 10). Compared to the associations with the AD converters group, we observed slightly more lipid species showing significantly altered trajectories in the combined AD- and MCI-converter group.

We selected several lipid species that showed differential trajectories as case studies to examine the trajectory over all time points (Figure 7C). AD and CN showed trajectories in opposite direction for several lipid species, including GM3(d18:1/24:1), Cer(d18:1/18:0), PE(P-16:0/22:6), and PI(18:0_22:6). By contrast, PC(O-16:0/16:0), AC(12:0), deDE(18:2), the oxidised lipid, PC(36:4) [+OH], the dimethyl-cholesteryl ester, dimethyl-CE(18:1), the bile acid dxCA, and CE(24:1) showed trajectories in the same direction. Interestingly, the trends of these lipid species in the converter group transitioned from CN concentrations at baseline to AD group concentrations by the end of the study.

### AD-related medications affect the trajectory of lipid species over time

The aforementioned, t-test results showed that anticholinesterase medication usage was significantly different between AD and CN groups. Further, longitudinal random forest backward selection analysis was performed to examine whether medications affect the trajectory of lipid species. The results showed that the trajectory of four lipid species deDE(18:2), deDE(20:4), GM3(d18:1/22:0), and PE(P)(18:1/22:4) were significantly (variable importance > 30) associated with these anti-dementia drugs (Supplementary Figure 4; Supplementary Table 11. Further, we also found that there were 91 and 160 lipid species (Supplementary Figure 4; Supplementary Table 11), whose trajectories were significantly affected by the medications of omega-3 and statins respectively.

### Trajectory of AD-CN scores varied among AD diagnosis groups

After evaluating the AD-CN model across all timepoints, we dervied the risk scores for each particiant. The distributions of AD-CN scores at each time point is shown in Supplementary Figure 5. We examined the association of the trajectories of the risk scores with AD status (non-converting CN, non AD-like MCI, AD-like MCI, AD converters, and AD) (Table 2). The cross-sectional associations of the lipidomic score with AD-like MCI versus non-converting CN (p=3.23×10^−26^) and AD converters versus the combination of non-converting CN and MCI groups (p=3.20×10^−16^) were signficant (Table 2). When we examined the changes of the overal AD-CN lipidomic scores among these groups, the AD-like MCI versus non-converting CN behaved signficantly different (p=3.78×10^−04^). The changes of AD converters and the non-converting CN and MCI groups were signficant different (p=3.27×10^−03^). The AD group behaved similarly as the CN group.

**Table 2.** The association of overall lipidomic scores with AD diagnosis (Intercept and trajectory).

| AD diagnosis groups | Associations in intercept |  |  |  | Associations in trajectory |  |  |  |
| --- | --- | --- | --- | --- | --- | --- | --- | --- |
|  | Beta * | 95% CI (Lower) | 95% CI (Upper) | P value | Beta# | 95% CI (Lower) | 95% CI (Upper) | P value |
| non AD-like MCI vs. non-converting CN | -0.192 | -0.306 | -0.077 | 1.07x10<sup>-03</sup> | 0.049 | 0.026 | 0.072 | 2.63x10<sup>-05</sup> |
| AD-like MCI vs. non-converting CN | 0.759 | 0.621 | 0.897 | 3.23x10<sup>-26</sup> | -0.046 | -0.071 | -0.021 | 3.78x10<sup>-04</sup> |
| AD vs. non-converting CN | 0.699 | 0.575 | 0.823 | 2.73x10<sup>-27</sup> | -0.003 | -0.033 | 0.026 | 8.39x10<sup>-01</sup> |
| AD converters vs. non-converters (CN+MCI) | 0.531 | 0.405 | 0.657 | 3.20x10<sup>-16</sup> | -0.026 | -0.044 | -0.009 | 3.27x10<sup>-03</sup> |

\*Mean difference in intercept.
#Mean change in slope over time.

The trajectory of the AD-CN score for all groups is shown in Figure 8. The trajectory of CN and AD groups showed minor trends in the oppostie direction. Compared to CN and AD groups, AD-like and non AD-like MCI showed much larger changes in oposing directions with the AD-like group showing a decrease and the non-AD like showing an increase in the AD-CN score. The trajectory of AD converters and non converters was also in oposing directions with the AD converters showing a slight decrease in the AD-CN score and the non-converters showing a slight increase.

**Figure 8.**
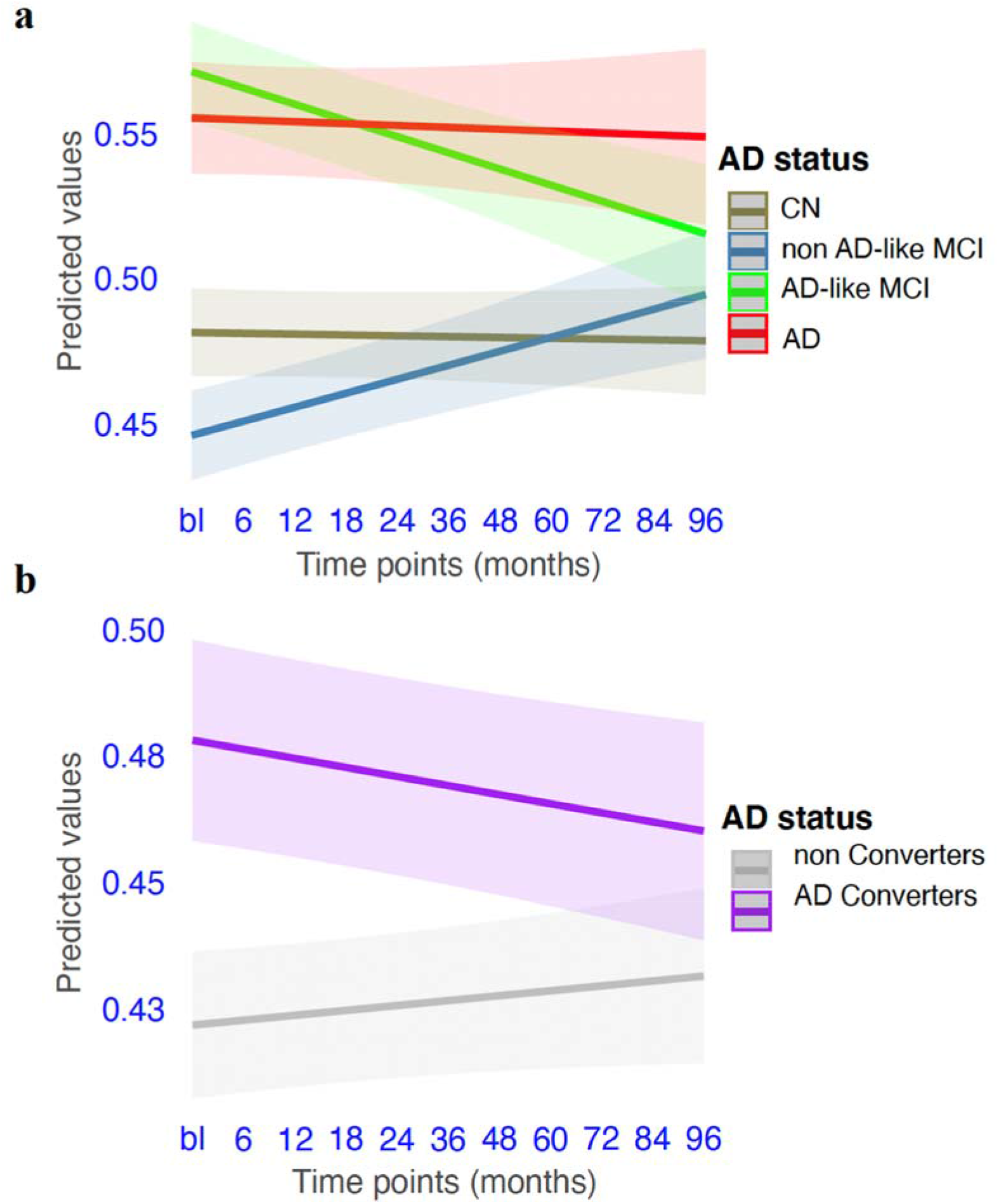
Trajectory of AD-CN scores between different AD diagnosis groups. **a**. The change of scores among non-converting CN, non AD-like MCI, AD-like MCI, and AD; **b**. The change of scores between AD converters and non converters (the combination of non-converting CN and MCIs). Linear mixed model was used to examine the changes of AD-CN scores among AD diagnosis groups adjusted by age, sex, BMI, fasting status, AD diagnosis status at baseline, total cholesterol, HDL-C, triglycerides, cohort, omega-3, and statin status.

## Discussion

We performed comprehensive cross-sectional and longitudinal analyses of lipidomic data in the ADNI cohort to identify the associations of the lipid species trajectories with AD diagnosis. Of note, there were many MCI participants in the ADNI study presenting with mixed aetiologies. While some progressed to AD over the follow-up period, others appeared to be stable but displayed heterogeneity in their plasma lipidome, and this was used to stratify these individuals into AD like and non-AD like MCI participants.

Previous research^5,6,29-36^ has also recognised this heterogeneity, with some studies implementing machine learning methods on neuroimaging data^29-34^ or plasma metabolites^35,36^ to predict MCI participants at high-risk of converting to AD. These studies showed promising results but were limited by small sample size. In this study, we developed an AD-CN lipidomic model, using ridge regression, within a 5-fold cross validation framework, on the large training dataset (AD and healthy groups; n=651). The prediction accuracy of the lipidomic model was assessed within the cross-validation framework of the training dataset and gave an AUC of 0.75 for the separation of CN and AD participants. Further, the AD-CN model efficiently classified MCIs (including AD converters) into low AD risk and high AD risk, with the high AD risk group having two times higher risk of conversion to AD than the low AD risk group. Using the model to stratify the non-converting MCI group showed 142 out of 413 MCI participants were defined as AD-like MCI (participants with high “lipidomic similarity” to AD).

The later trajectory analysis of lipid species between AD-like and non AD-like MCI groups delineate the heterogeneity of the MCI groups. We also observed that the AD-like MCI group had a significantly larger proportion of individuals taking anticholinesterase inhibitors, indicating that our AD-CN classification model could efficiently capture individuals in the MCI group who had started to develop AD symptoms.

The metabotype conservation index showed that overall, the lipidome was stable across time (the first two years). However, the conservation index in clinical AD was significantly lower than the CN group indicating greater variation in the lipidome over the two years of this analysis. While AD converters and the non-converting MCI group were not significantly different to the CN group.

In contrast to the conservation index calculated over two years, the lipid trajectories (calculated over 10 years) showed multiple lipid species with trajectories that associated with AD converters relative to the non-converting CN and MCI groups. We identified a greater decrease of (LPC(O), LPC(P)), LPC, and PC(O) species in the converter group. In particular, PC(O-38:5) and PC(O-40:5) decreased during the progression to AD. There was consistent evidence of decreasing ether lipids for participants in the transition to AD^28^, which may reflect changes in the biosynthetic pathway i.e. a gradual deterioration in peroxisome function, leading to decreased ether lipids in circulation^37^. AD converters also showed a strong decrease in lysophospholipids species (containing 16:0, 18:0, 18:1, 20:0, 22:0, 22:1 and 24:0 fatty acids) during the progression to AD. In support of this observation, several cross-sectional studies have reported that plasma levels of LPC were decreased in the AD patients compared with healthy group^38^ and the LPC-to-PC ratio were also inversely associated with AD^39,40^, suggesting decreasing phospholipase activity as the disease progresses. In humans, LPC(O) and LPC(P) are metabolised by both these metabolic pathways – 1) these lipids are synthesised as ether lipids, originating in the peroxisomes; and 2) these lipids are produced by a plasmalogen specific phospholipase A2 (PLA2), which cleaves the fatty acid from the sn-2 position. Therefore, we hypothesise that both pathways – peroxisome dysfunction^37^ and decreased PLA2^41,42^ – result in the scarcity of lysophospholipids in participants transitioning to AD. A better understanding of the altered activity of PLA2 and ether lipid metabolism could help to identify novel therapeutic targets in AD^43^.

The lipidomic score provides a global view of the changes in the lipidome over time among AD diagnosis groups. The lipidomic score for non-converting CN and AD groups were stable across time, with AD showing a higher level than the CN group. Consistent with the differences in the trajectories of the lyso and ether lipid species observed between AD converters and non converters (Figure 7b), the overall lipidomic score between the two groups showed significant changes in opposite directions (Figure 8b). Further, we observed differences in the lipidomic score between the AD-like and non AD-like MCI groups and this agreed with the greater number of individual lipid species trajectories showing a significant difference between these groups (Figure 7a). Perhaps surprisingly the AD-like MCI group showed a downward trend in the lipidomic score towards the CN value, suggesting the lipidome may be normalising in this group, while the non AD-like MCI group showed an increase in the lipidomic score crossing the CN value. Clearly these two groups represent different metabolic phenotypes with the AD-like MCI being stable and resilient to progression to AD over the 10 year follow up period. This may be associated with a normalisation of their metabolic phenotype as measured by the AD-CN score. In contrast, the non-AD like group which starts with a low AD-CN score appears to progress as evidenced by an increasing AD-CN score but not to the point of conversion (Figure 8a). These global and individual lipid species trajectories may provide useful biomarkers to monitor disease progression and better target MCI participants at greatest risk of progression to AD for clinical trials or treatment. However, further studies are required to define the outcomes for the AD-like and non AD-like metabolic phenotypes identified in this study.

There were several limitations in this study. Although the follow-up time of the ADNI study extends to 10 years, the majority of records are from baseline to 24 months. The metabotype conservation index under two years framework has limited ability to capture the variations of each individual among different AD diagnosis groups. And, the power of the longitudinal analysis on the cohorts within a 2-year period is limited. In addition, a second cohort is needed to externally validate the performance of our AD-CN classification model.

In conclusion, we have performed comprehensive lipidomic analyses using the longitudinal ADNI -1, -2 and -GO cohorts. At baseline, we have developed a novel AD-CN classification model to characterise the heterogeneity of MCI participants, providing the potential of using lipidomics to efficiently distinguish high-risk subjects within the MCI group. The subsequent longitudinal analysis using the data set across all the time points highlighted significant changes in the lipidome over time in AD converters relative to CN and AD-like MCI relative to non AD-like MCI groups. These highlight the potential of lipidomic studies to improved our understanding of the relationships between lipid metabolism and progression to AD. Lipidomic biomarkers also show promise to improve clinical risk assessment and management of older individuals at risk of AD.

## Methods

### Participants

Alzheimer’s Disease Neuroimaging Initiative (ADNI)-1, -2 and -GO (http://adni.loni.usc.edu/) is a longitudinal study, recruiting 1,517 individuals over 55 years old at baseline. At intervals of 6-12 months, blood and clinical data were collected from each individual, up to a maximum of 10 years. Lipidomic profiling was performed on all blood samples, with 4,873 plasma samples examined from baseline up to the 13^th^ time point (at 10 years followed up). After filtering 143 missingness, we ended up with 4,730 samples in total. At baseline, there were originally 1,418 individuals, of which we excluded 25 individuals with missing values (10 missing cognitive scores, 3 missing fasting information, and 12 missing BMI). Thereby, there remained 1,393 participants at baseline in the study.

The definition of probable AD in ADNI followed the NINDS-ADRDA criteria^44^. In brief, individuals with Mini-Mental State Exam (MMSE) scores between 20 and 26 (inclusive) and a Clinical Dementia Rating Scale (CDR) of 0.5 or 1.0 were classified as AD patients^45^. Participants were defined as MCI if they had MMSE scores between 24 and 30, a memory complaint, objective memory loss measured by education-adjusted scores on Wechsler Memory Scale Logical Memory II, a CDR of 0.5, absence of significant levels of impairment in other cognitive domains, and essentially preserved activities of daily living^46^.

We further defined the longitudinal status of AD diagnosis group. At baseline, there were 437 CN, 713 MCI, and 243 AD cases. Among all these 1,393 participants, we defined two categories according to their changes of diagnosis status over time: 1) AD converters – 329 participants whose status were CN/MCI at baseline but converted to AD later; 2) MCI converters – 71 CN progressed to MCI and stayed as MCI at the following time points. Additionally, we also observed there were 53 MCI reverted back to CN. And, there were 39 individuals (10 CN, 5 AD, and 24 MCI at baseline) whose status varied across time points. We treated these unstable individuals as undefined (might be affected by the medication usages) and removed them from the longitudinal analysis (though in our cross-section analysis such as AD-CN modelling, we still keep them in the analysis).

### Lipidomic profiling

Lipidomic profiling was performed on all plasma samples (n=4,730) using our recently expanded, targeted lipidomic profiling strategy using reverse phase liquid chromatography coupled to an Agilent 6495C QqQ mass spectrometer. The lipid extraction and LC-MS/MS methodology, using scheduled multiple reaction monitoring (MRM), was as previously described^15^ with the addition of approximately 200 novel lipid species from 17 lipid classes^24^. Further details about our latest methodology to measure these lipids are described on our laboratory website (https://metabolomics.baker.edu.au/method/).

Overall, there were 781 lipid species from 49 lipid classes quantified. Single ion monitoring (SIM) and neutral loss (NL) are two types of measurements for the same TG lipid species, with NL measurements more specific and sensitive. To avoid the redundancy and improve the accuracy in our modelling analysis, we excluded 32 TG[SIM] lipids, retaining 749 lipid species from 48 lipid classes in this study. The details for all the lipid species and classes were listed in Supplementary Table 1.

### Statistical analysis

In the following analysis, log10 transformation followed by a standard normalisation (zero mean and one-unit standard deviation) was performed on individual lipid species across repeated measurement of all participants. We introduced the covariate set including age, sex, BMI, HDL-C, total cholesterol, triglycerides, fasting status, cohort (a categorical variable indicating ADNI 1, GO, and 2 phases), omega-3, and statin status for the following models.

### Development of an AD-CN model

We sought to use the normalised lipidomic data on AD subjects (n=243) and cognitive normal individuals (CN; n=408) at baseline to build the classification model. Further, we applied the model to stratify non-converting MCI (n=413) from AD converters (n=329). Ridge regression models were created to stratify AD from CN, optimizing C-statistic using the R package ‘glmnet v4.1-4’. Five models were created from an external 5-fold cross-validation framework (Figure 1). All these models were adjusted for age, sex, BMI, *APOE ε*4, HDL-C, total cholesterol, triglycerides, fasting status, cohort (a categorical variable indicating ADNI 1, GO, and 2 phases), omega-3, and statin status. Since we identified that two deDE lipid species (deDE(18:2) and deDE(20:4)) were strongly associated with dementia-related medication, in the models, we used the whole lipidomes except the two deDE lipid species as the main predictors.

Beta coefficients from each cross-validation fold were averaged. This model was then applied to the non-converting MCI group to generate the probabilities of the MCI individuals being “AD-like” or “non AD-like”.

In addition, the weights were separately applied to the whole dataset across different time points to generate overall AD risk scores for each individual at each time.

### The metabotype conservation index

As changes in metabolic phenotypes (metabotype) over time can be indications for disease onset or progression^47^, we used the metabotype conservation index described by Yousri et al.^48^ to quantify the stability of the metabotype of ADNI participants over time. This analysis was restricted to comparisons between baseline and follow up visits after 12 and 24 months, which had 755 common participants with samples across baseline, 12 month and 24 months.

As pairwise correlation-based metrics can be distorted by the strong correlation structure observed for lipid measurements, we first aggregated strongly correlated lipids into representative variables (“eigenlipids”). To this end, we first clustered lipid concentrations at baseline using weighted correlation network analysis using the R packages WGCNA (version=1.71)^49^ and dynamicTreeCut (version=1.63.1)^50^. We used a soft threshold of 0.86, which was closest to the recommended threshold of 0.9 ^51^ and corresponded to a power of 10 used for the calculation of the adjacency matrix. We then calculated the topological overlap matrix (TOM) using the adjacency matrix. The resulting distance matrix was used for hierarchical clustering and the dendrogram was cut using the hybrid method of the cutTreeDynamic-function using the following parameters: deepSplit=3, pamDendroRespect=TRUE, and minClusterSize = 2. The latter was chosen to sensitively extract clusters that capture all strong correlations down to at least two lipids. We then calculated one eigenlipid for each individual cluster by extracting its first principal component. Metabolites in the outgroup were retained as separate features. We additionally provide results for several other values of the parameter minClusterSize (5, 10, and the default value of 20). Cluster assignments for each lipid across settings is provided in Supplementary Table 12, with results for higher values of the minimum cluster size (5, 10, and 20) in Supplementary Figure 6 and Supplementary Table 3.

Projecting the cluster and outgroup assignment from baseline to 12 months and 24 months, we obtained longitudinal measures to calculate the metabotype conservation index *L*_*c*_*(x)* for subject *x* as

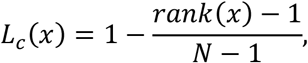

where *rank(x) = N – z* and 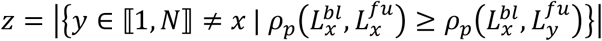 with *N* being the number of subjects, 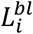 being the aggregated lipidomics profile of subject *i* at baseline, 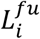 being the aggregated lipidomics profile of subject *i* at follow up, and 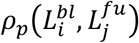 being the Pearson correlation coefficient of the lipidomics profile of subject *i* at baseline and the lipidomics profile of subject *j* at follow up for *x, i, j ∈* ⟦1,*N*⟧. The resulting *L*_*c*_*(x)* ranges between 0 and 1, with 0 meaning no conservation and 1 meaning maximal conservation. We compared proportions of individuals with *I*_*c*_ = 1 and *I*_*c*_ < 1 per diagnostic group using Fisher’s exact test, considering the stable CN group as reference. The resulting distribution of *I*_*c*_ values smaller than 1 (i.e., individuals whose lipid profiles change over the two-year period) were compared across diagnostic groups using the Wilcoxon rank sum test. For assessing changes in the *I*_*c*_ distribution globally between baseline to 12 months and baseline to 24 months, we used both paired t-tests and the Wilcoxon test.

#### Longitudinal analysis using linear mixed models

We calculated linear mixed models using repeated measurements across 13 time points to examine the associations between AD diagnosis state and trajectory of lipid species (either species level or overall lipidomic scores) over time using the interaction between time points and AD state. In the model, we treated normalised individual lipid species or overall lipidomic scores as the independent variables, and AD diagnosis state (the category variable) as the main predictor and adjusted the models with a list of covariates of age (at baseline), sex, BMI, HDL-C, total cholesterol, triglycerides, fasting status, cohort, time point (treated as continuous variable), omega-3, and statin status. The interaction between time point and AD diagnosis was introduced into the model, which is the key term for examining the trajectory of lipid species over time among AD states. In the model, we perform two sets of longitudinal analyses on different subsets of the population to: 1) examine the difference in the trajectory of lipid species among AD patients, CN, and non-converting MCI group (AD-like or non AD-like) using the whole population (excluding the converters); 2) use the changes in lipid species to predict the AD converters on the population excluding all prevalent AD cases. The lme4 package in software R 3.6.2 was used to perform the linear mixed models.

#### Sensitivity analysiss

We performed sensitivity analysis to examine whether the cross-sectional associations between AD state (AD cases vs CN) and lipid species are consistent across the major time points (baseline, 12 months, and 24 months). To perform this, we used linear regression with lipid species as the independent variable, and AD state as the main predictor.

To assess whether anticholinesterases were associated with specific lipid trajectories, random forest backward selection^52^ was performed using the Boruta function implemented within the Boruta R package (version 7.0.0). The calculated predictive value (termed as variable importance, calculated across 100 permutations of the original dataset) reflects the strength of the association between medications and the changes in lipid species. Predictive values of medication (grouped by ATC codes) on lipids with importance larger than 30 (mean + 4 standard deviations) were chosen as highly relevant^52^.

We performed another sensitivity analysis to evaluate whether the trajectories of lipid species within MCI converters behaved similarly as those within AD converters. To do this, linear mixed models was carried out to examine the associations of trajectory of lipid species with the combination of MCI converters and AD converters relative to non-converting CN and MCI groups.

## Data Availability

The results published here are in whole or in part based on data obtained from the AD Knowledge Portal. ADNI associated data is also available from the Laboratory of Neuro Imaging Image and Data Archive at https://ida.loni.usc.edu/login.jsp.

## Acknowledgement

Support was provided by the National Health and Medical Research Council of Australia (# 1101320 and 1157607). KH is supported by an NHMRC Investigator grant (GNT1197190). This work was also supported in part by the Victorian Government’s Operational Infrastructure Support Program.

The data available in the AD Knowledge Portal would not be possible without the participation of research volunteers and the contribution of data by collaborating researchers.

Data collection and sharing for this project was funded by the Alzheimer’s Disease Neuroimaging Initiative (ADNI) (National Institutes of Health Grant U01 AG024904) and DOD ADNI (Department of Defense award number W81XWH-12-2-0012). ADNI is funded by the National Institute on Aging, the National Institute of Biomedical Imaging and Bioengineering, and through generous contributions from the following: AbbVie, Alzheimer’s Association; Alzheimer’s Drug Discovery Foundation; Araclon Biotech; BioClinica, Inc.; Biogen; Bristol-Myers Squibb Company; CereSpir, Inc.; Cogstate; Eisai Inc.; Elan Pharmaceuticals, Inc.; Eli Lilly and Company; EuroImmun; F. Hoffmann-La Roche Ltd and its affiliated company Genentech, Inc.; Fujirebio; GE Healthcare; IXICO Ltd.; Janssen Alzheimer Immunotherapy Research & Development, LLC.; Johnson & Johnson Pharmaceutical Research & Development LLC.; Lumosity; Lundbeck; Merck & Co., Inc.; Meso Scale Diagnostics, LLC.; NeuroRx Research; Neurotrack Technologies; Novartis Pharmaceuticals Corporation; Pfizer Inc.; Piramal Imaging; Servier; Takeda Pharmaceutical Company; and Transition Therapeutics. The Canadian Institutes of Health Research is providing funds to support ADNI clinical sites in Canada. Private sector contributions are facilitated by the Foundation for the National Institutes of Health (www.fnih.org). The grantee organization is the Northern California Institute for Research and Education, and the study is coordinated by the Alzheimer’s Therapeutic Research Institute at the University of Southern California. ADNI data are disseminated by the Laboratory for Neuro Imaging at the University of Southern California.

Metabolomics data is provided by the Alzheimer’s Disease Metabolomics Consortium (ADMC) and funded wholly or in part by the following grants and supplements thereto: NIA R01AG046171, RF1AG051550, RF1AG057452, R01AG059093, RF1AG058942, U01AG061359, U19AG063744 and FNIH: #DAOU16AMPA awarded to Dr. Kaddurah-Daouk at Duke University in partnership with a large number of academic institutions. As such, the investigators within the ADMC, not listed specifically in this publication’s author’s list, provided data along with its pre-processing and prepared it for analysis, but did not participate in analysis or writing of this manuscript. A complete listing of ADMC investigators can be found at: https://sites.duke.edu/adnimetab/team/.

Data on lipidome was generated at Baker Heart and Diabetes Institute, a member of ADMC. Details on the lipid profiling technologies are described at: (https://metabolomics.baker.edu.au/method).

## Author Contributions

Meikle, Kaddurah-Daouk and Kastenmüller led the study design team. Wang, Arnold, and Huynh led the statistical analyses presented in this study. Giles and Livera provided statistical suggestions on linear mixed models. Weinisch and Marella performed the analysis on metabotype conservation index and random forest backward selection. Mellett, Duong, Huynh and Giles supported the acquisition and processing of the lipidomic data for the cohort. Arnold, Kastenmüller, Nho, Saykin, Han and Kaddurah-Daouk were key members of the ADNI team and represent the Alzheimer’s Disease Metabolomics Consortium (ADMC): A complete listing of ADMC investigators can be found at https://sites.duke.edu/adnimetab/who-we-are/.

