## Supplementary Figures for "Trajectory of plasma lipidomes associated with the risk of late-onset Alzheimer’s disease pathogenesis: a longitudinal study in the ADNI cohort"

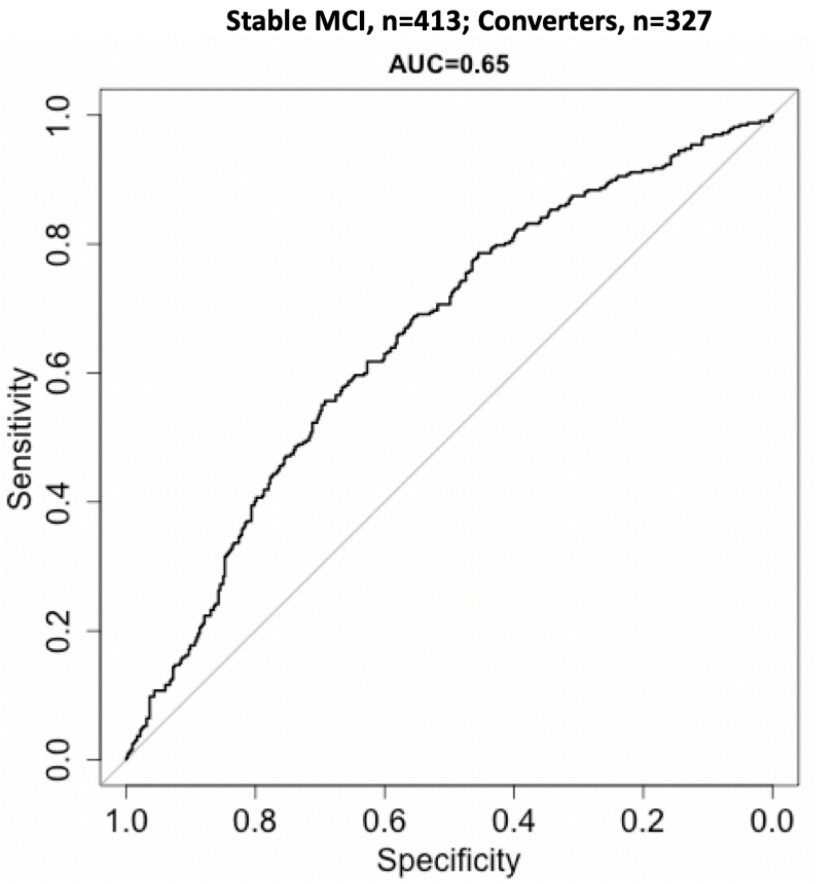


**Supplementary Figure 1. The prediction performance of AD-CN model using the basic predictor set including age, sex, BMI and *APOE ε*4.**

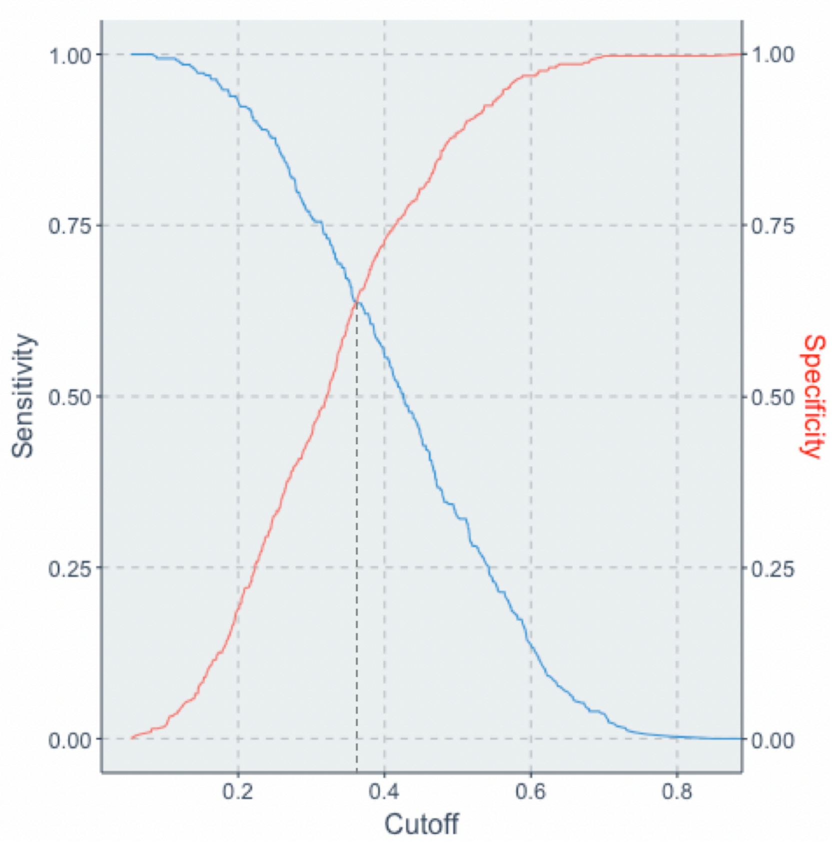


**Supplementary Figure 2. The definition of cut-off point (predicted value=0.37) to stratify the converters-MCI groups into AD-like and non AD-like.**

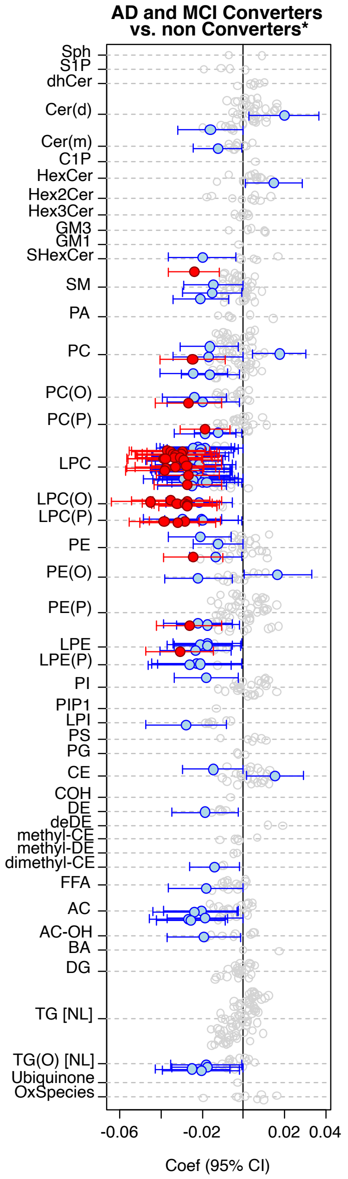


**Supplemental Figure 3. Trajectory of lipid species between the non Converters (CN and MCI) and the combination of AD and MCI converter.**

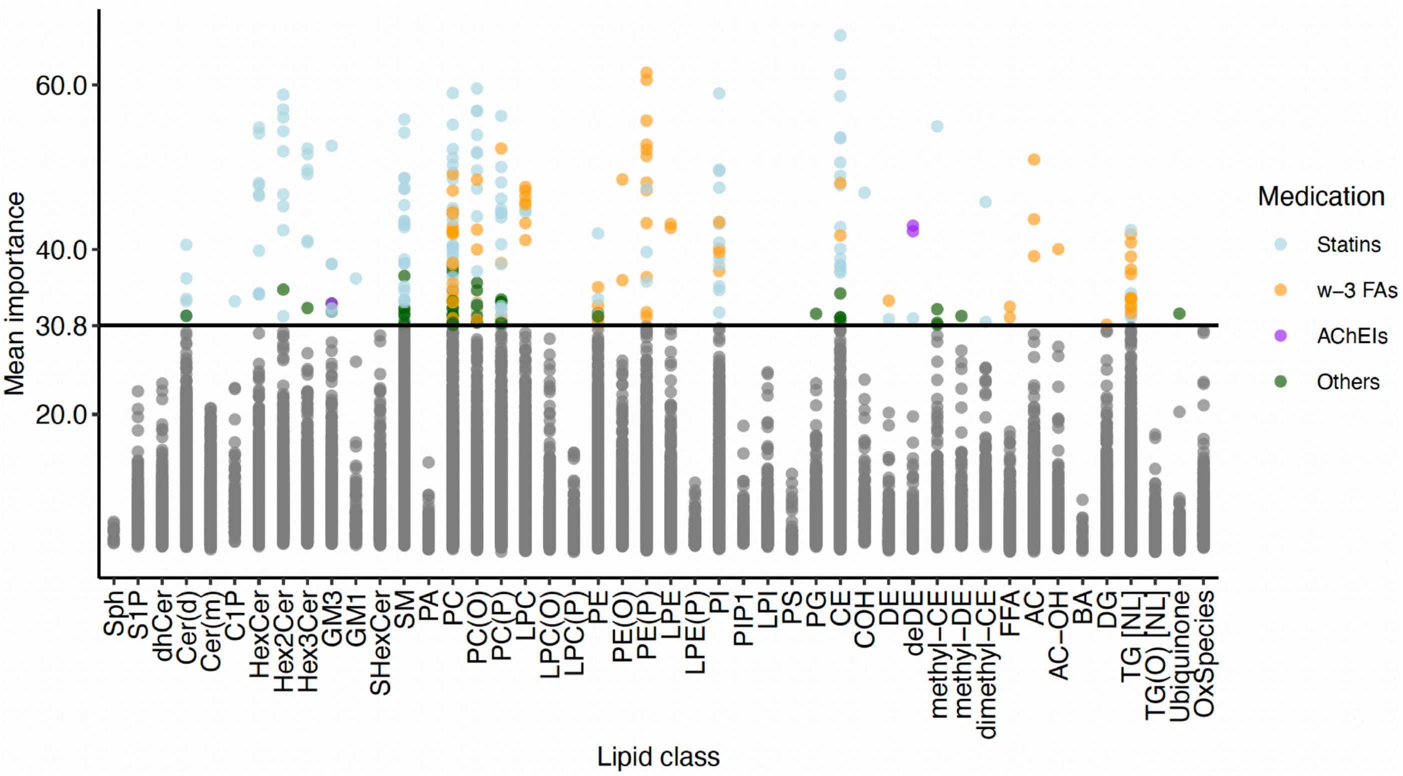


**Supplemental Figure 4. Feature selection of significant medication effects using longitudinal random forests.** Medications with effects on lipids (grouped by pathway) are highlighted based on their mean importance, which represents the strength of association of medications with a lipid species. This importance is calculated based on the dataset containing all lipid species measured at month 0, month 12, and month 24 using longitudinal random forests. Medication with a mean importance higher than mean+4sd across importance values are considered significant. Each dot represents the predictive value of a single medication onto a single lipid. We observed a strong overrepresentation of statins and omega-3 fatty acid (w-3 FAs) supplementation, while other medication classes showed only few effects on the lipidome. Anticholinesterases (AChEIs) are highlighted as a special case, given their highly significant and selective effects on two deDe lipid species, which seems to be an unknown off-target side effect of this class of anti-dementia medications.

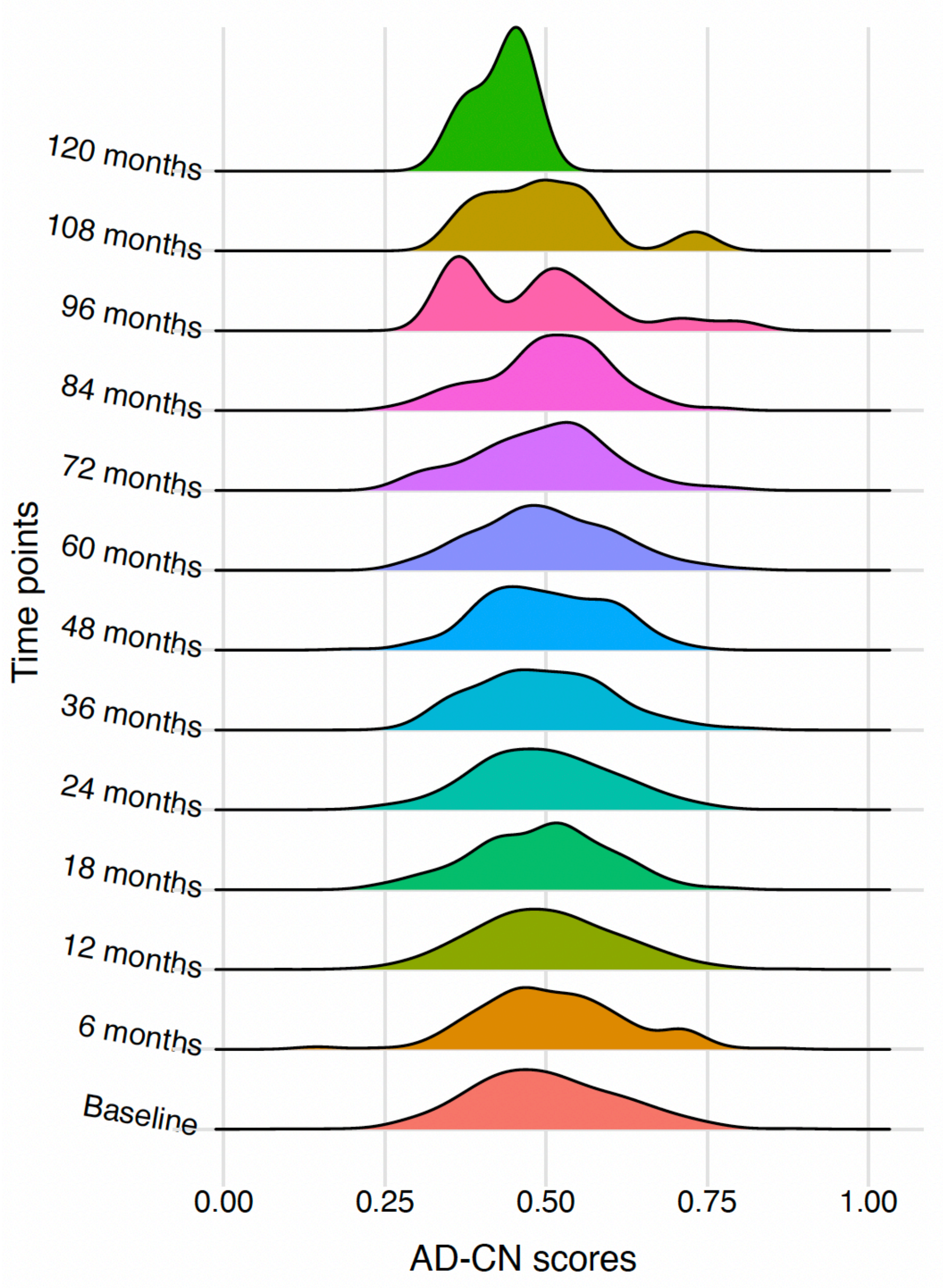


**Supplemental Figure 5. The distribution of the derived AD-CN scores across all the time points.**

The lipidomic scores on each single time point were calculated using the weights from ridge regression models on AD-CN data set at baseline.

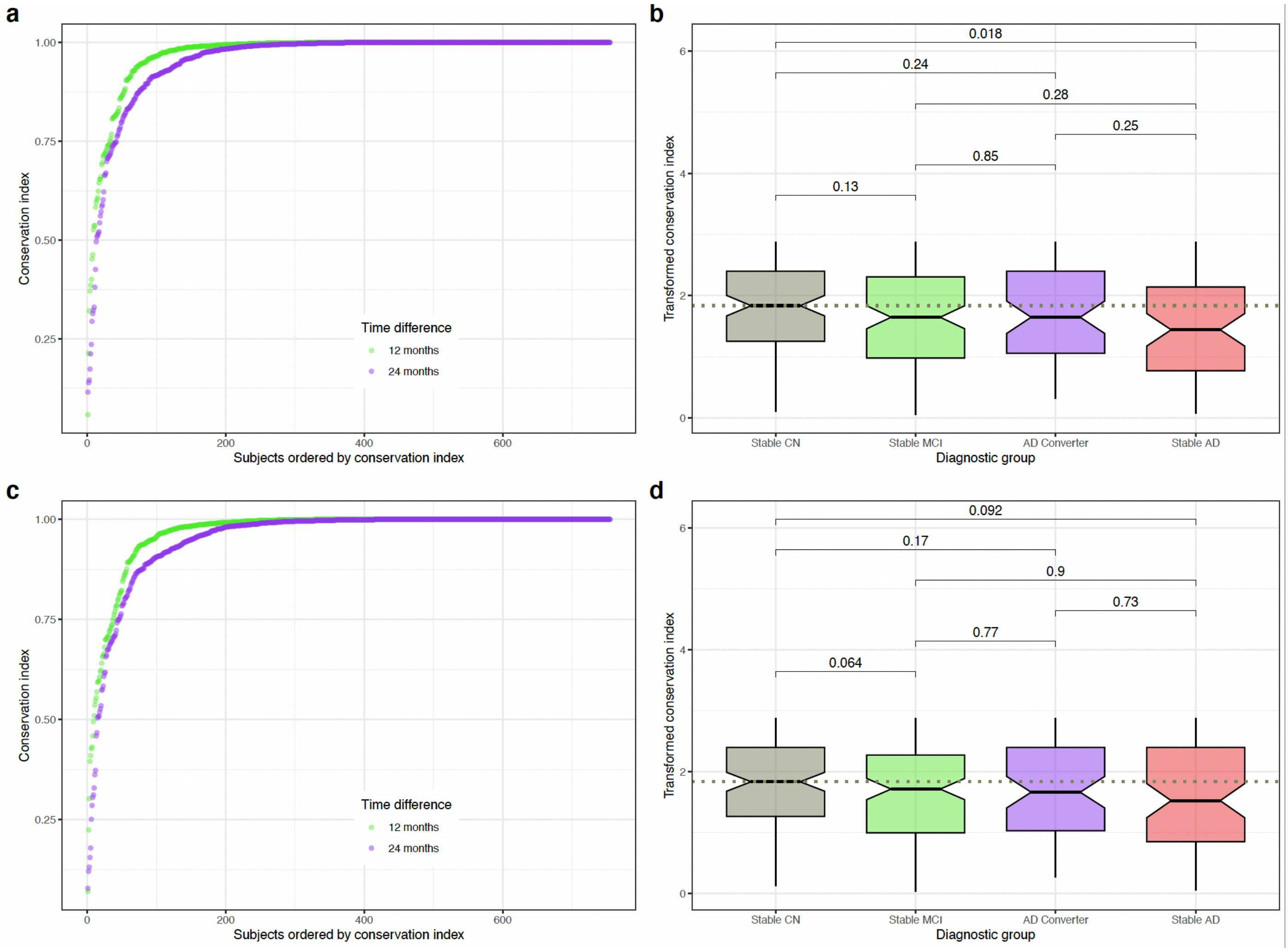


**Supplementary Figure 6. Findings for the metabotype conservation index analysis for different minimum cluster sizes used for calculating eigenlipids. a.** and **b.** showing results for mininmum cluster size=5. **c.** and **d.** showing results for minimum cluster size=10. **e.** and **f.** showing results for the default minimum cluster size=20.
